# Impact of Synchronized Left Ventricular Pacing in Cardiac Resynchronization Therapy

**DOI:** 10.1101/2023.02.27.23286538

**Authors:** Yuichiro Miyazaki, Kohei Ishibashi, Nobuhiko Ueda, Satoshi Oka, Akinori Wakamiya, Keiko Shimamoto, Kenzaburo Nakajima, Tsukasa Kamakura, Mitsuru Wada, Yuko Inoue, Koji Miyamoto, Satoshi Nagase, Takeshi Aiba, Kengo Kusano

**Author notes:** **Correspondence:** Kohei Ishibashi, MD, PhD, Department of Cardiovascular Medicine, National Cerebral and Cardiovascular Center, 6-1, Kishibe-Shimmachi, Suita, Osaka 564-8565, Japan.

## Abstract

**Background:** The adaptive cardiac resynchronization therapy (aCRT) algorithm enables synchronized left ventricular pacing (sLVP) to produce fusion with intrinsic right ventricular activation in normal atrioventricular (AV) conduction. Although sLVP presents benefits over biventricular pacing (BVP), the adequate sLVP rate for better clinical outcomes remains unclear. This study aimed to assess the association between sLVP rates and clinical outcomes.

**Methods:** Among our cohort of 271 consecutive patients who underwent CRT implantation between April 2016 and August 2021, we evaluated 63 patients who underwent CRT without considerably prolonged AV conduction and applied the aCRT algorithm (48 men, mean age: 64 ± 14 years; median follow-up period: 316 days [interquartile range: 212–809 days]).

**Results:** At the 6-month follow-up after CRT implantation, the frequency of CRT responders was 71% (n = 45). The sLVP rate was significantly higher in responders than in non-responders (75 ± 30 vs. 47 ± 40 %, *p* = 0.003). Receiver operating characteristics (ROC) curve analysis revealed that the optimal cut-off value during the sLVP rate was 59.4% for prediction of CRT responders (area under the curve, 0.70; sensitivity, 80%; specificity, 61%; positive predictive value, 84%; and negative predictive value, 55%). Kaplan–Meier analysis demonstrated that the higher sLVP group (sLVP ≥59.4%, n = 43) had better prognosis (cardiac death and heart failure hospitalization) than the lower sLVP group (sLVP <59.4%, n = 20) (log-rank *p*<0.001), and multivariate Cox hazard analysis revealed that a higher sLVP rate was associated with good prognosis (*p*<0.001).

**Conclusion:** sLVP was associated with CRT response, and higher sLVP rate (≥59.4%) is important for good prognosis in patients with aCRT.

**CLINICAL PERSPECTIVE:** *What Is New?:*

- Synchronized left ventricular pacing(sLVP) >59.4% was a significant predictor of cardiac resynchronization therapy(CRT) responders and better clinical outcomes, evidenced by the results of the multivariate analysis.
- In this study, which included patients with moderately prolonged PR intervals, high sLVP rates were associated with better clinical outcomes.

*What Are the Clinical Implications?:*

- sLVP rate was associated with the improvement of cardiac function after CRT implantation.
- A higher sLVP rate was associated with a lower risk of cardiac death and heart failure hospitalization.

## 1 INTRODUCTION

Cardiac resynchronization therapy (CRT) improves cardiac function and clinical outcomes in patients with symptomatic heart failure accompanied by decreased left ventricular ejection fraction (LVEF), QRS prolongation, and left bundle branch block (LBBB) (1). However, up to 30% of patients do not see improvements in cardiac function and/or clinical prognosis after CRT implantation (2). The Adaptive CRT algorithm (aCRT) is a novel pacing algorithm for CRT that continuously optimises the atrioventricular (AV) and the interventricular (VV) delays based on heart rate and intrinsic AV conduction (3). In cases whereby the AV interval is shorter than 220 ms (during atrial sensing) or 270 ms (during atrial pacing), the aCRT algorithm uses the RV intrinsic conduction, which can provide synchronized left ventricular pacing (sLVP) to create fusion beat with the intrinsic conduction. In cases where the AV interval is extremely prolonged, the aCRT pacing provides biventricular pacing (BVP) without fusion with the intrinsic conduction. The sLVP was reported to have benefits over BVP regarding the risk of heart failure (HF), hospitalization, cardiac death, or atrial fibrillation (4,5). However, the adequate sLVP rate for better clinical outcomes remains unclear. This novel study aimed to assess the appropriate sLVP rate for recovering cardiac function in the mid-term, and to improve clinical outcomes in the long-term.

## 2 METHODS

### 2.1 Study population

This retrospective single-centre study included all consecutive patients who underwent CRT device implantation featuring the Adaptive CRT algorithm (Medtronic Inc., Minneapolis, MN, USA) (aCRT) between April 1, 2016, and August 31, 2021. The aCRT pacing mode was adapted just after the implantation. Patients who applied the pacing algorithm without adopting sLVP mode were excluded from this analysis. In addition, patients with persistent atrial fibrillation, AV block, and extremely prolonged PR also were excluded from this analysis, due to difficulty in using sLVP.

This study was conducted in accordance with the principles outlined in the Declaration of Helsinki and approved by the ethics committee of our institution (M26-150-10). This is a retrospective study to analyze the anonymous data generated after patients have agreed to treatment; we applied the opt-out method to obtain informed consent.

### 2.2 Study protocol

The patients enrolled in this study were classified into two groups (CRT responder or non-responder) depending on the response to CRT pacing 6 months after CRT implantation. Next, we determined the required sLVP rate to become a CRT responder using a receiver operating characteristic (ROC) curve analysis. Thereafter, we compared the association between the sLVP rate and clinical prognosis.

### 2.3 Cardiac examinations and definition of responses before and after CRT implantation

Cardiac systolic function was assessed using both echocardiography and single-photon emission computed tomography (SPECT) before and 6 months after CRT device implantation. Electrocardiogram (ECG)-gated myocardial perfusion SPECT imaging with technetium-99m (^99m^Tc) sestamibi was performed to measure cardiac systolic function and dimensions. All echocardiographic data were measured during three consecutive cardiac cycles. The left ventricular end-systolic volume (LVESV) and LVEF were measured using Simpson’s biplane method.

CRT responders were described as patients with an improved LVEF >10% and/or reduced LVESV >15% 6 months after CRT device implantation compared with baseline. Cardiac death and/or hospitalization for HF were evaluated as clinical outcomes. Cardiac death was defined as HF death, left ventricular assist device implantation, or death from ventricular arrhythmia. HF hospitalization was defined as a sudden or gradual onset of symptoms of New York Heart Association (NYHA) functional class III or IV HF, requiring unplanned hospitalization.

### 2.4 Statistical analyses

Data were analyzed using JMP software (version 11.2.01, SAS Institute, Cary, NC, USA). Continuous variables are presented as mean ± standard deviation and were compared using Student’s *t*-test. Categorical variables were compared using Fisher’s exact test. The cumulative incidence and event-free curves were based on Kaplan–Meier analyses, stratified by study group, and contrasted using the log-rank test. Hazard ratios and odds ratios are reported with their 95% confidence intervals. To identify independent predictors of clinical response, univariable analyses were first performed, and predictors with a significance level <5%, age, and sex were included in the multivariable models. Multivariable analyses were performed using a Cox regression model. *P*-values of <0.05 were considered reflective of statistically significant differences.

## 3 RESULTS

### 3.1 Patient characteristics

Among our cohort of 271 consecutive patients who underwent CRT implantation between April 2016 and August 2021, we evaluated 63 patients who underwent CRT without considerably prolonged AV conduction on whom we applied the aCRT algorithm (48 men, mean age: 64 ± 14 years) (Figure 1). The baseline characteristics of the patients are shown in Table 1. The number of patients with ischaemic cardiomyopathy (ICM) was 17 (27%), and mean LVEF was 23 ± 7%. Mean PR and QRS time were 192 ± 32 ms and 155 ± 25 ms, respectively, and the number of patients with LBBB was 31 (49%).

**Table 1.**
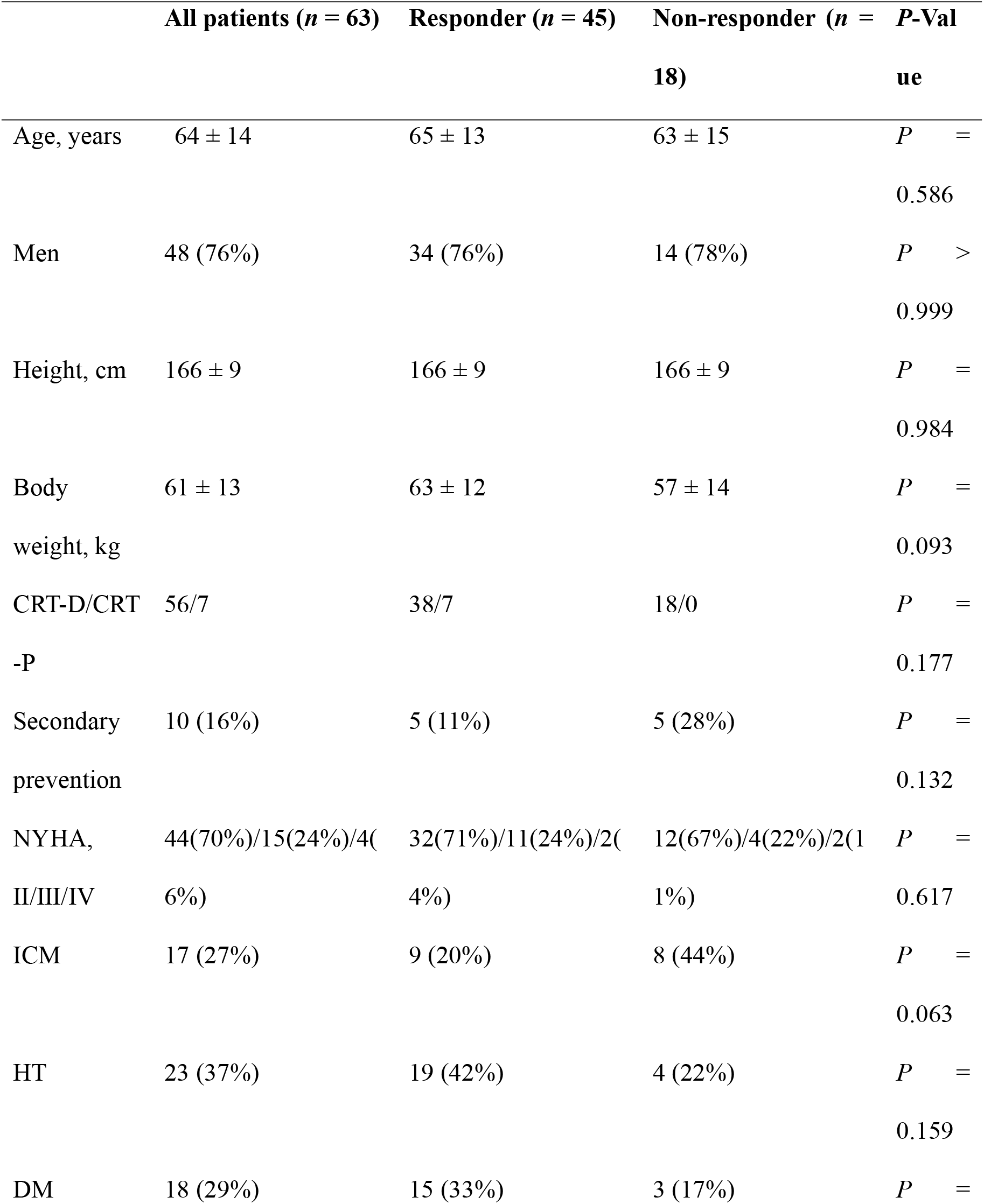
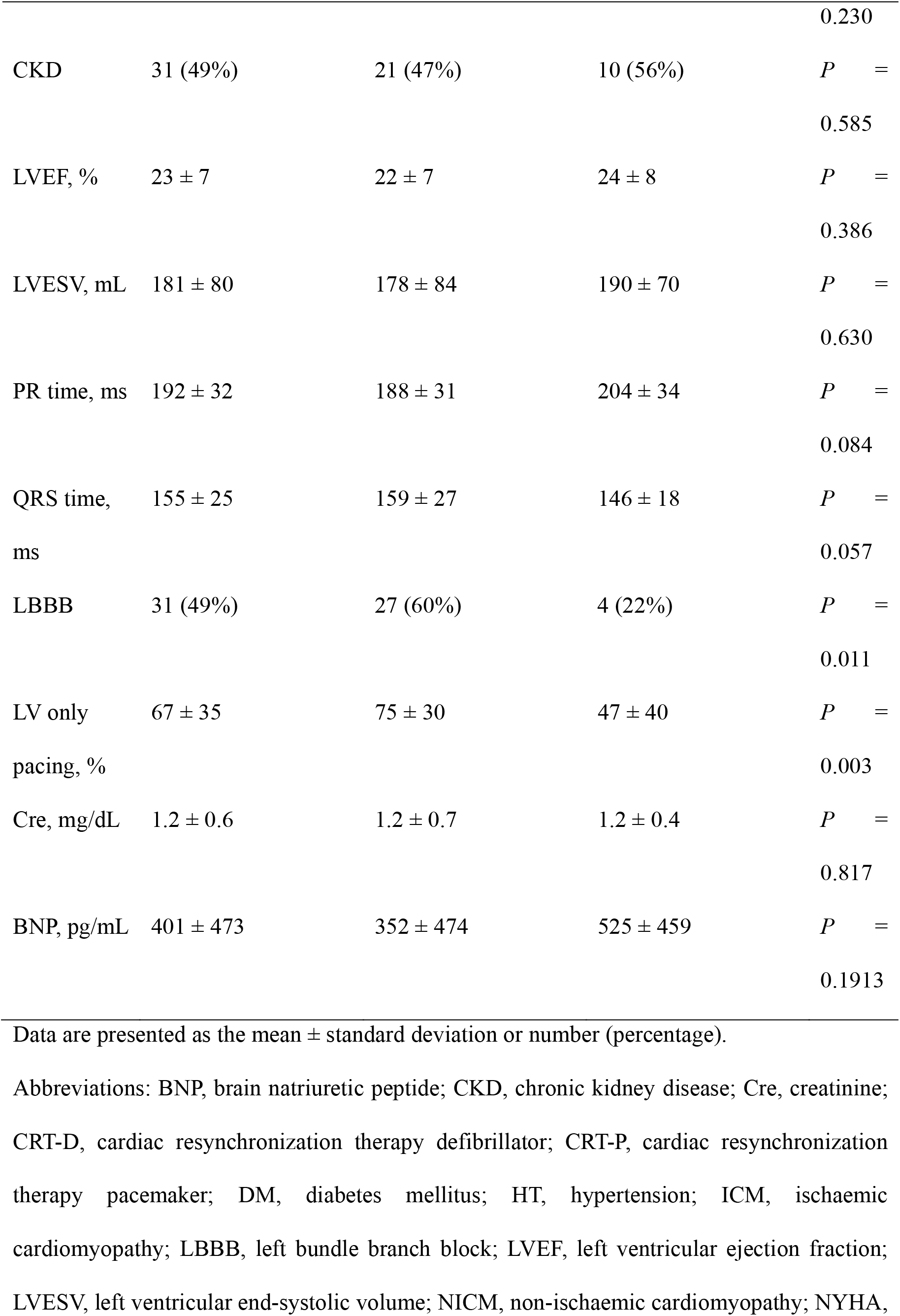
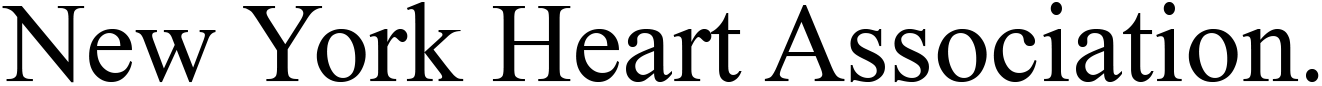
Baseline characteristics between responders and non-responders.

**Figure 1.**
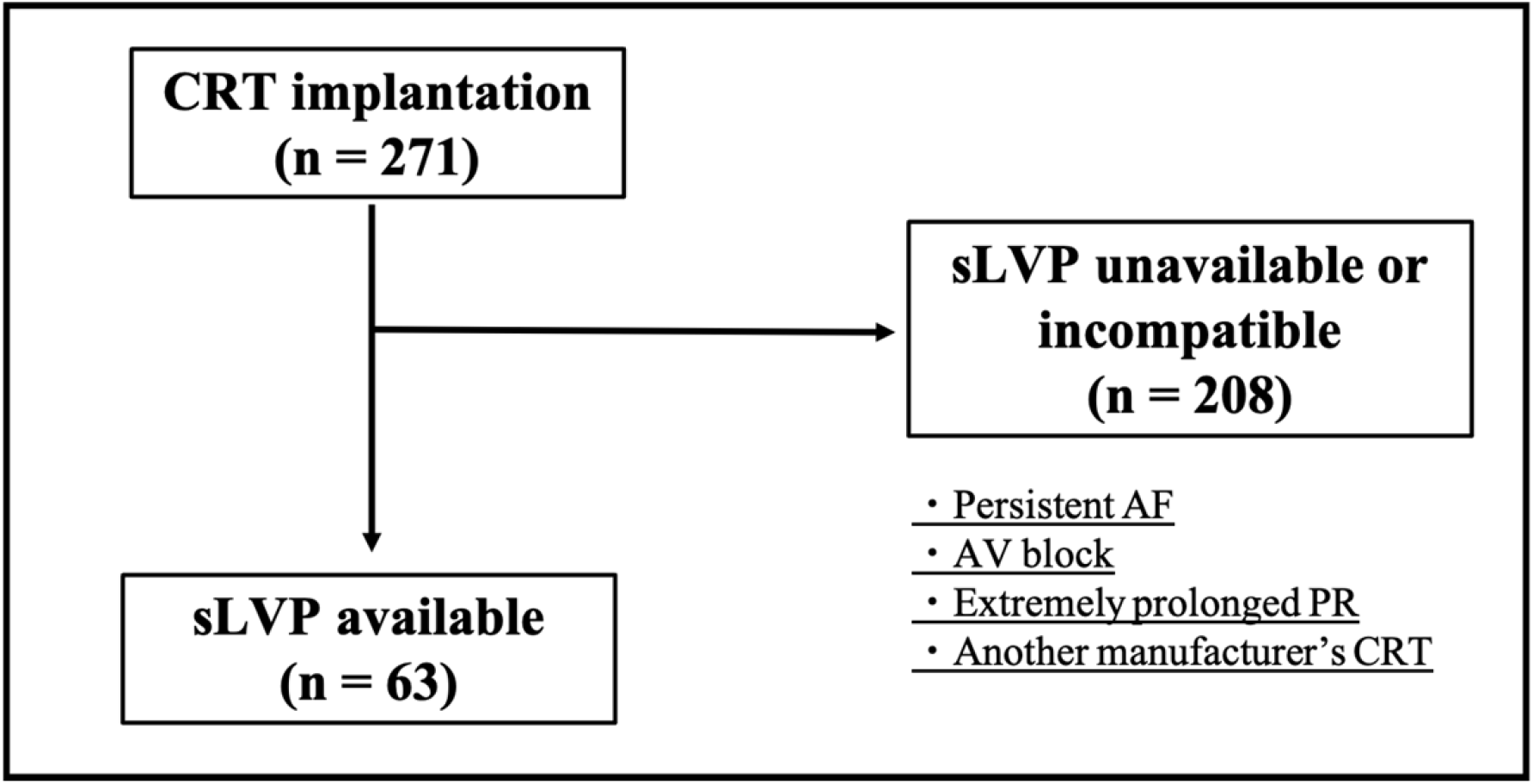
Overview of the study population. Among our cohort of 271 consecutive patients who underwent CRT implantation between April 2016 and August 2021, we evaluated 63 sLPV enabled CRT patients and applied the aCRT algorithm. Abbreviations: AF, atrial fibrillation; AV, atrioventricular; CRT, cardiac resynchronization therapy; sLVP, synchronized left ventricular pacing.

### 3.2 CRT responder and synchronized left ventricular pacing rate

At the 6-month follow-up after CRT implantation, the frequency of CRT responders was 71% (n = 45) (Table 1). Patients with LBBB were significantly more frequent than those without. There was no significant difference in the QRS width and rate of ICM. The sLVP rate was significantly higher in responders than in non-responders (75 ± 30 vs. 47 ± 40%, *p* = 0.003). ROC curve analysis revealed that the optimal cut-off value during sLVP rate was 59.4% for the prediction of CRT responders (area under the curve, 0.71; sensitivity, 80%; specificity, 61%; positive predictive value, 84%; and negative predictive value, 55%) (Figure 2). Multivariate analysis showed that sLVP >59.4% was an independent responder for CRT responder (odds ratio: 4.52, 95% CI: 1.18–18.90, *p* = 0.027) (Table 2).

**Table 2.**
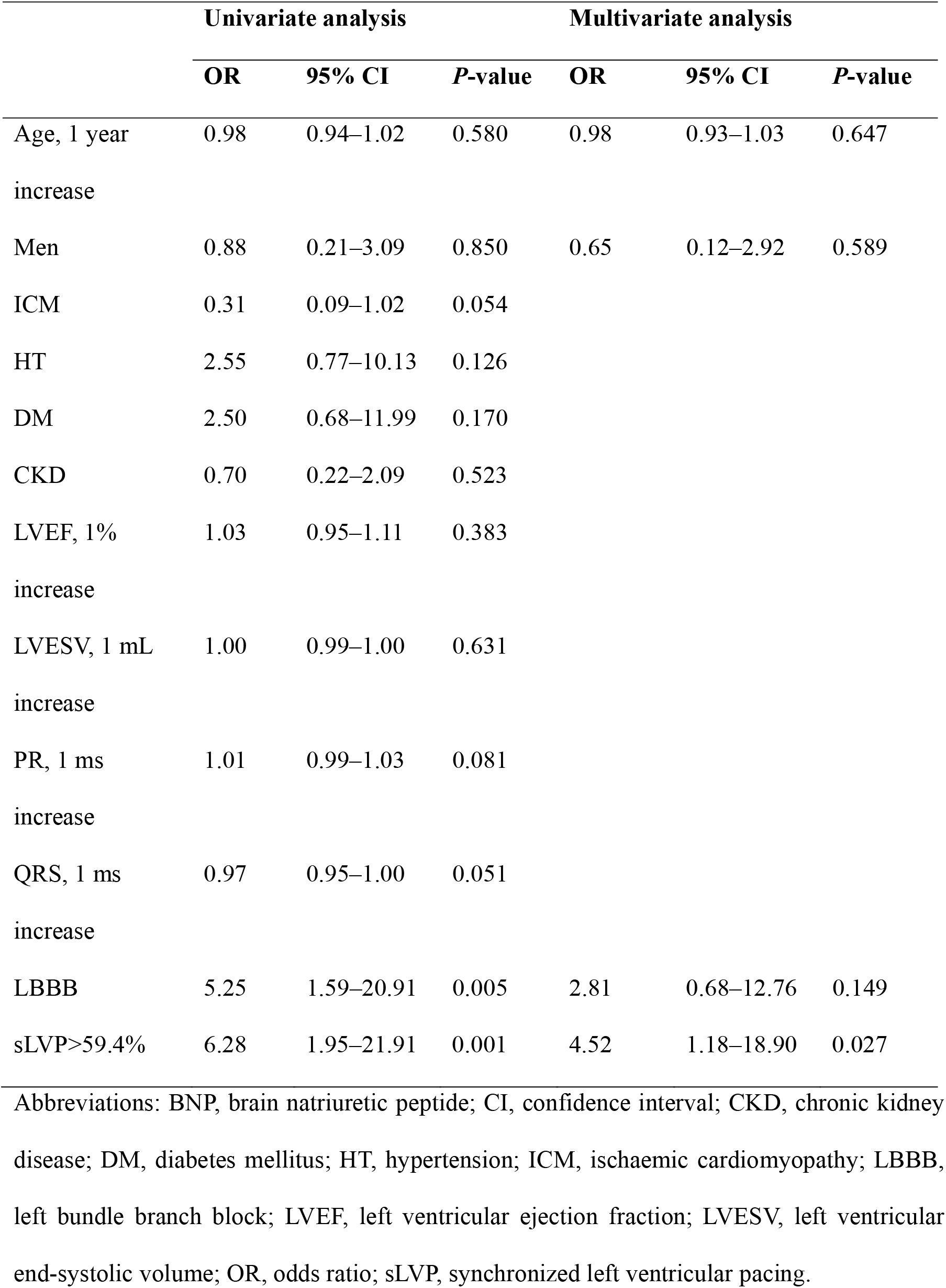
Univariate and multivariate analyses for responder.

**Figure 2.**
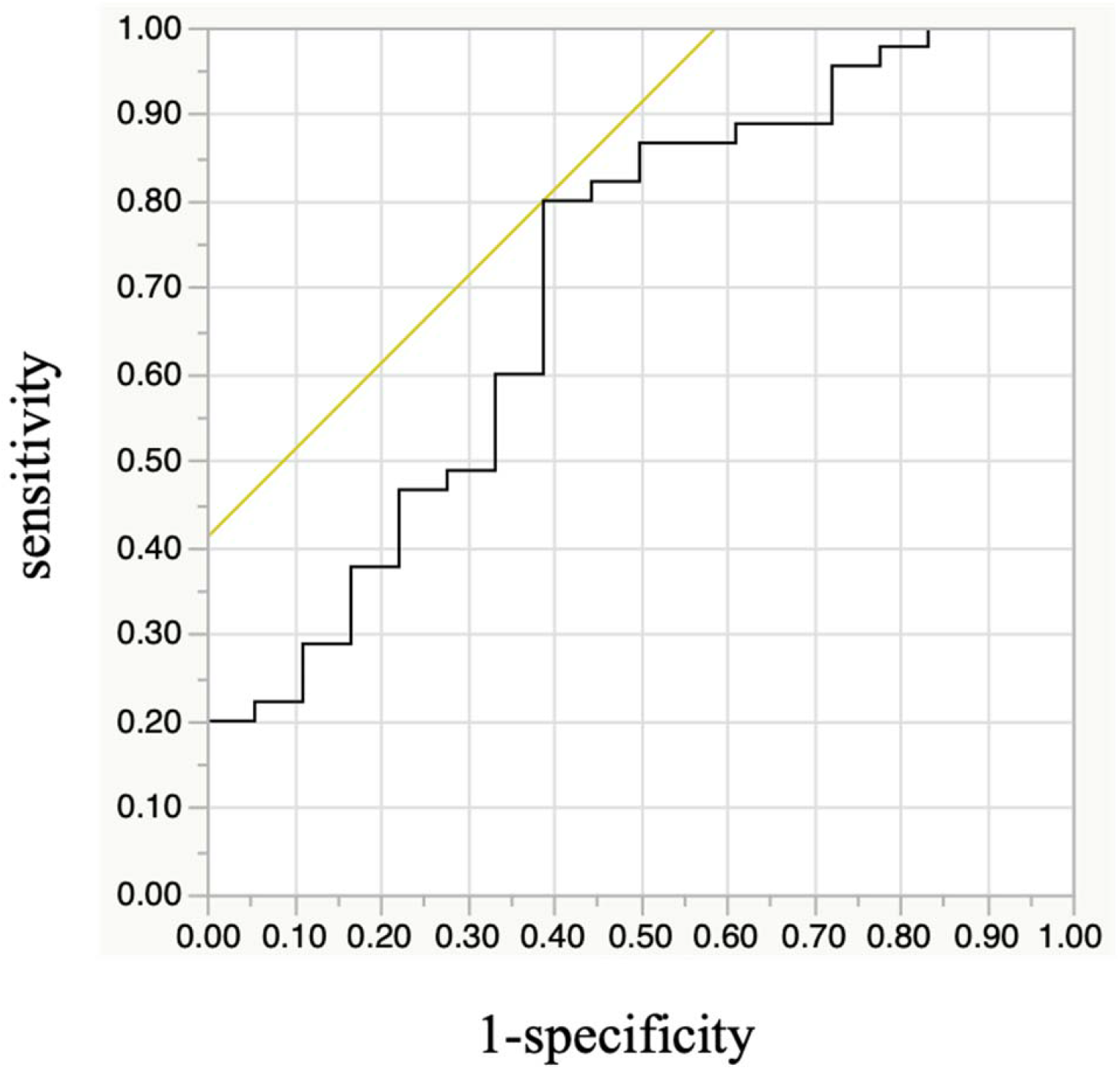
The association between CRT responder and synchronized left ventricular pacing rate. Receiver operating characteristic curve analysis revealed that the optimal cut-off value during sLVP rate was 59.4% for the prediction of CRT responders. Abbreviations: CRT, cardiac resynchronization therapy; sLVP, synchronized left ventricular pacing

### 3.3 Clinical outcome of high sLVP rate

During a median follow-up of 346 days (IQR, 223–760 days), one patient had cardiac death and 13 patients had HF hospitalizations. Five patients died during hospitalization because of HF. Kaplan–Meier analysis demonstrated that the high sLVP group (sLVP ≥59.4%, n = 43) had a better prognosis (cardiac death and HF hospitalization) than the low sLVP group (sLVP <59.4%, n = 20) (log-rank *p*<0.001) (Figure 3), and multivariate Cox hazard analysis revealed that higher sLVP rate was associated with good prognosis (hazard ratio: 0.10, 95% CI: 0.01– 0.40, *p* = 0.027, *p*<0.001) (Table 3).

**Table 3.** Univariate and multivariate analyses for cardiac death and heart failure hospitalization.

|  | Univariate analysis |  |  | Multivariate analysis |  |  |
| --- | --- | --- | --- | --- | --- | --- |
|  | HR | 95% CI | *P*-value | HR | 95% CI | *P*-value |
| Age, 1 year increase | 0.99 | 0.96–1.04 | 0.972 | 0.95 | 0.90–1.00 | 0.061 |
| Men | 0.68 | 0.22–2.53 | 0.541 | 0.63 | 0.15–2.88 | 0.541 |
| ICM | 2.51 | 0.82–7.32 | 0.102 |  |  |  |
| HT | 0.28 | 0.04–1.03 | 0.055 |  |  |  |
| DM | 1.00 | 0.27–3.04 | 0.999 |  |  |  |
| CKD | 3.72 | 1.23–13.72 | 0.019 | 14.51 | 2.79–108.45 | <0.001 |
| LVEF, 1% increase | 0.97 | 0.89–1.04 | 0.521 |  |  |  |
| LVESV, 1 mL increase | 1.00 | 0.99–1.00 | 0.994 |  |  |  |
| PR, 1 ms increase | 1.01 | 0.99–1.03 | 0.060 |  |  |  |
| QRS, 1 ms increase | 0.98 | 0.96–1.00 | 0.149 |  |  |  |
| LBBB | 0.26 | 0.05–0.84 | 0.023 | 0.41 | 0.08–1.55 | 0.198 |
| sLVP>59.4% | 0.12 | 0.02–0.40 | <0.001 | 0.10 | 0.01–0.40 | <0.001 |

Abbreviations: CI, confidence interval; CKD, chronic kidney disease; DM, diabetes mellitus; HR, hazard ratio; HT, hypertension; ICM, ischaemic cardiomyopathy; LBBB, left bundle branch block; LVEF, left ventricular ejection fraction; LVESV, left ventricular end-systolic volume; sLVP, synchronized left ventricular pacing.

**Figure 3.**
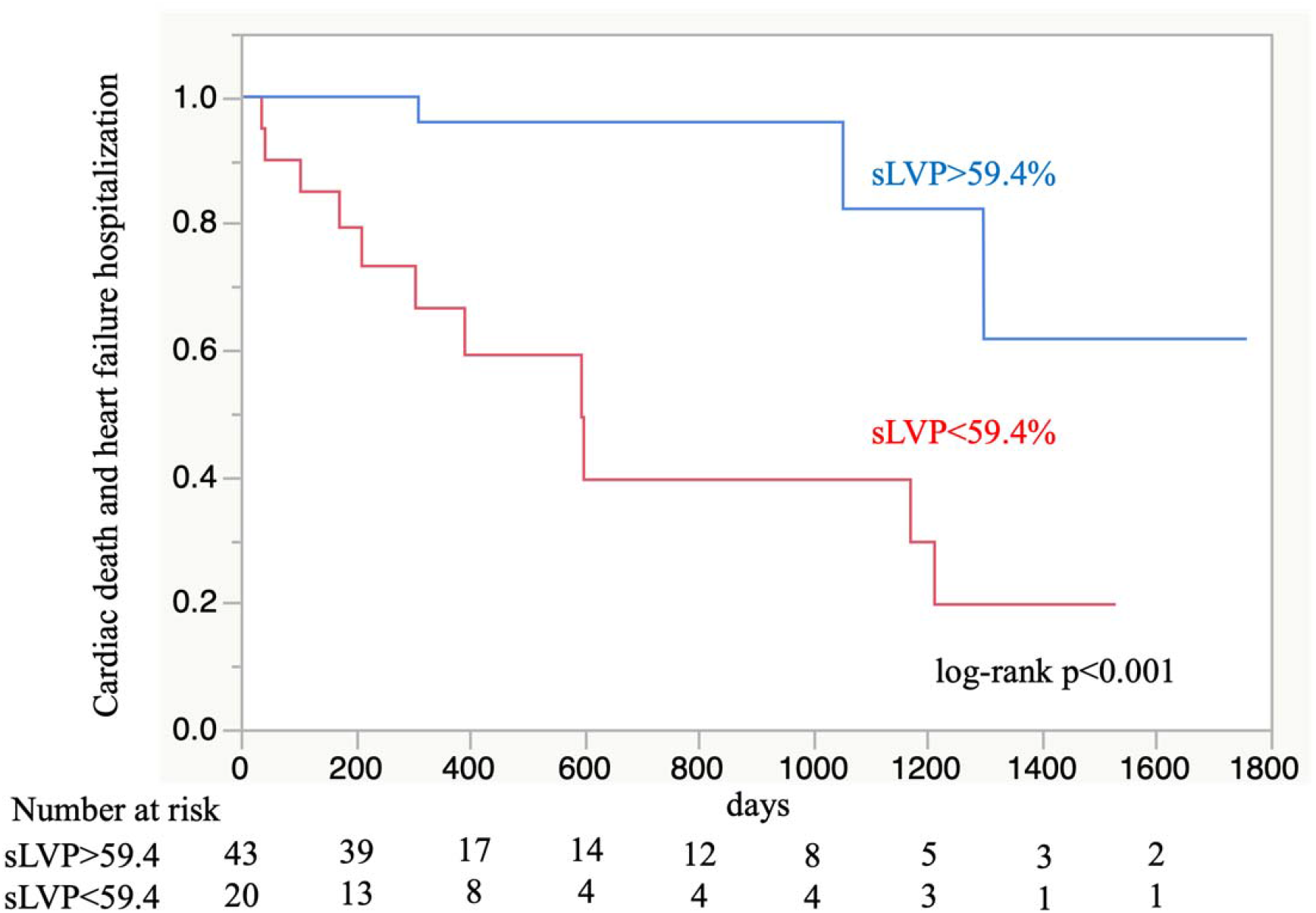
Kaplan–Meier analysis. The analysis demonstrates that the higher sLVP group (sLVP>59.4%, n = 43) had better prognosis (with regard to cardiac death and heart failure hospitalization) than the lower sLVP group (sLVP<59.4%, n = 20) (log-rank *p*<0.001). Abbreviations: sLVP, synchronized left ventricular pacing.

## 4 DISCUSSION

### Main findings

The main findings of this study are as follows: (1) CRT responders had higher sLVP rates than CRT non-responders, (2) sLVP >59.4% was an excellent predictor of CRT response, and (3) sLVP >59.4% led to fewer cardiac deaths and hospitalizations due to HF.

### The mechanism of fusion beat of pacing and intrinsic conduction

Recent CRT devices have the function to automatically adjust the fusion beat of self AV conduction and pacing. This regulatory function seems to lead to further narrowing of the QRS width after the CRT implantation (6). Previous studies have reported that the narrowing of QRS width after the CRT implantation can improve echocardiographic outcomes and long-term mortality (7,8). In addition to the function of adjusting the fusion beat, other studies have reported that sLVP could shorten QRS width compared with BVP (9,10). The sLPV may contribute to improved responder rate and better clinical outcomes by achieving QRS narrowing by creating the fusion beat with the intrinsic waveform and pacing.

### The adequate frequency of synchronized left ventricular pacing

Adaptive CRT has an automatic adjustment algorithm for AV and interventricular delays based on frequent AV conduction evaluations. Furthermore, sLVP is often used for normal AV conduction (9). A previous study showed that sLVP provided better clinical outcomes than conventional BVP (4); however, the frequency of sLVP that leads to a better prognosis is not well known. Previous studies have defined the cut-off rate of sLVP to be 80% to predict clinical and echocardiographic parameters (9). Another study reported that an sLVP rate >50% was associated with better clinical outcomes compared with that of <50% (4). However, these rates are not statistically determined values. In this study, sLVP >59.4% was a significant predictor of CRT responders and better clinical outcomes, evidenced by the results of the multivariate analysis. Using this criterion, we may be able to predict clinical responses, such as cardiac death and HF hospitalization.

### The association between synchronized left ventricular pacing and PR interval

A previous study reported that LVP improved LV dp/dt max in patients with a PR interval <200 ms (11). Other studies have also reported the effect of sLVP in patients with a normal PR interval (9,10); however, the significance of sLVP in patients with prolonged PR intervals is unknown. In this study, which included patients with moderately prolonged PR intervals, high sLVP rates were associated with better clinical outcomes. However, the patients with sLVP <59.4% had longer PR intervals than the patients with sLVP >59.4%. The sLVP rate appeared significantly affected by PR interval, especially in patients with prolonged PR intervals. Prolonged PR intervals are considered a significant factor in preventing a high sLVP rate. A previous report showed that RA septal pacing was associated with a shorter AV interval than right appendage pacing (12). The case report demonstrated that the sLVP rate was increased by RA septal pacing in patients with extremely prolonged PR intervals (424 ms). In this study, most patients experienced right appendage pacing. We should consider that the RA pacing lead is positioned at the RA septum in patients with a prolonged PR interval to increase the LPV rate.

## 5 STUDY LIMITATIONS

This study had some limitations. First, this was a retrospective single-centre study. The results of this study should be further confirmed by prospective and multi-centre studies in the future. Second, the sample size was small, which reduces the statistical strength of our findings. In order to resolve this point, it is necessary to collect cases in a multi-center study. Third, the current study compared the mixture of sLVP and BVP. To clarify the effect of the sLVP, we have to compare the pure sLVP and the pure BVP.

## 6 CONCLUSIONS

Our study shows that the sLVP rate is associated with the CRT responder rate. A higher sLVP rate (>59.4 %) is associated with a lower risk of cardiac death and HF hospitalization.

## Data Availability

The datasets generated and/or analyzed during the current study are available from the corresponding author upon reasonable request.

## LIST OF ABBREVIATIONS

aCRT: Adaptive cardiac resynchronization therapy
AV: Atrioventricular
BVP: Biventricular pacing
ECG: Electrocardiogram
HF: Heart failure
ICM: Ischaemic cardiomyopathy
LBBB: Left bundle branch block
LVEF: Left ventricular ejection fraction
LVESV: Left ventricular end-systolic volume
NYHA: New York Heart Association
ROC: Receiver operating characteristics
SPECT: Single-photon emission computed tomography
sLVP: Synchronized left ventricular pacing
VV: Interventricular

